# Patients’ accounts of living with and managing Inflammatory Bowel Disease in rural Southern New Zealand: a qualitative study

**DOI:** 10.1101/2020.06.25.20133421

**Authors:** Lauralie Richard, Geoff Noller, Sarah Derrett, Trudy Sullivan, Fiona Doolan-Noble, Andrew McCombie, Michael Schultz, Christine Ho, Tim Stokes

**Affiliations:** Department of General Practice and Rural Health, Dunedin School of Medicine, University of Otago, New Zealand; Department of Preventive and Social Medicine, Dunedin School of Medicine, University of Otago, New Zealand; Department of Medicine, Dunedin School of Medicine, University of Otago, New Zealand; Department of Gastroenterology, Southern District Health Board, Dunedin, New Zealand

## Abstract

**Objective:** To explore how adults living with Inflammatory Bowel Disease (IBD) in rural New Zealand manage their condition and engage with health care providers.

**Design:** Qualitative exploratory design with semi-structured interviews analysed thematically.

**Setting and participants:** Interviews were conducted with 18 people living with IBD in the Otago region of the South Island.

**Results:** Five important constructs were identified: 1. Journey to confirming and accepting diagnosis; 2. Importance of the relationship with the health care team; 3. Support from others; 4. Learning how to manage IBD; and 5. Care at a distance - experiences of rurality. Pathways to confirming diagnosis involved two contrasting journeys: a long and slow process where diagnosis remained unclear for a prolonged period, and a more acute process where diagnosis typically came as a shock. Central to the acceptance process was acknowledging the chronicity of the condition, which involved feelings of grief but also the fear of judgement and stigma. Building a strong relationship with the specialist was central to medical management, particularly in the initial stage following diagnosis. Support from others was critical, enabling participants to progress through acceptance of the disease and developing confidence in its everyday management. Participants shared different strategies on how to manage IBD, describing a “trial and error” process of “finding what’s right” at different stages of the condition. Managing IBD rurally involved challenges of access to specialist care, with perceptions of delayed referrals and concerns about disparities in specialist access compared to urban counterparts. Rural living also had financial implications - cost of time and cost of mobilising resources for long travels to the urban centre for treatments.

**Conclusions:** Findings from this study provide a rich understanding of the complex health journeys of people living with IBD and the challenges of managing the condition rurally.

**STRENGTHS AND LIMITATIONS OF THIS STUDY:** 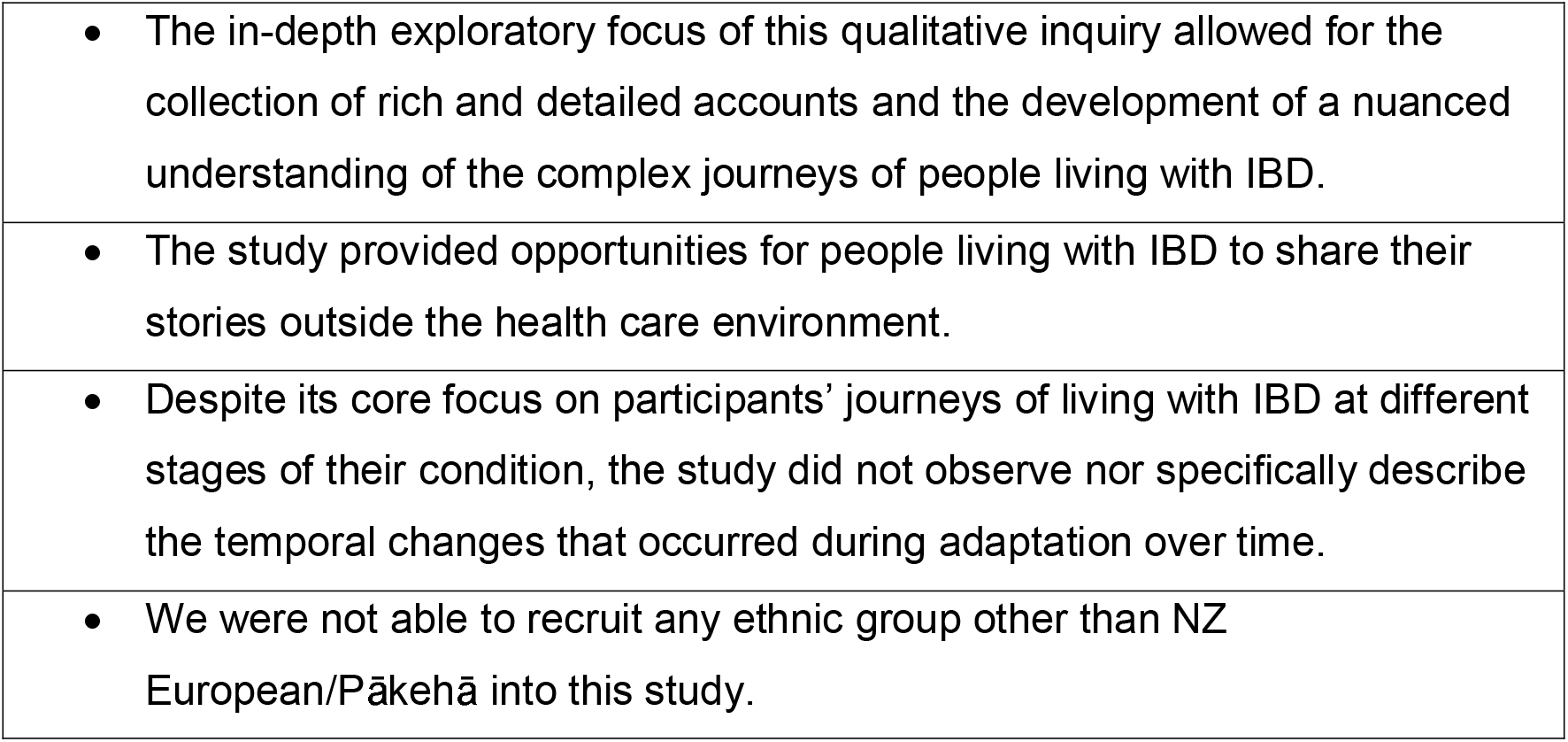

## INTRODUCTION

Inflammatory Bowel Disease (IBD) is a chronic inflammatory condition of the gastrointestinal tract which has three main subgroups: Crohn’s Disease, Ulcerative Colitis and IBD unclassified (IBDU).^1^ The prevalence of IBD exceeds 0.3% in North America, Oceania, and many European countries^2^, and the incidence and prevalence are increasing worldwide. New Zealand (NZ) has a relatively high incidence (30/100,000 in 2012) and prevalence of IBD in its NZ European (Pākehā) population; incidence and prevalence are lower in Māori, the indigenous people of NZ.^3^ IBD is a condition with considerable morbidity, having a chronic course alternating between active disease requiring urgent medical attention and periods of remission with routine health monitoring. Patients with active disease may experience very frequent bowel motions, often bloody, associated with faecal incontinence and urgency.^4^ Treatment aims to achieve both remission of symptoms, prevent disease flares and subsequent development of complications. Depending on disease severity this can be achieved through use of conventional medical therapy (e.g., amino-salicylates, immunomodulators), biological therapy and surgery.^5 6^

In NZ, as in other high income countries, people with IBD are managed by medical specialists (gastroenterologists) based in tertiary hospitals in urban centres and attend regular hospital based review appointments.^7^ Such a model of care may disadvantage those living rurally^7 8^ where considerable barriers may be faced in terms of timely access to health care, the need to travel long distances for specialist consultations, and the lack of appropriate resources for IBD management in their local community.^8 9^ This has led to the increasing use of new and alternative methods of delivering gastroenterology care to rural IBD patients such as telehealth, online support, and remote outreach clinics.^10-12^

There is a growing body of qualitative research evidence that explores patients’ experiences of living with and managing their IBD.^4 13^ Patients are particularly affected by the physical symptoms of IBD, which have an adverse effect on their psychological and social well-being. This literature is, however, limited in its exploration of the support patients receive from their health care providers.^4 13^

This study aimed to explore how adults living with IBD in rural NZ manage their condition and engage with their health care providers.

## METHODS

### Study setting

This research took place in the Central Otago and Queenstown-Lakes Districts of the Otago Region, in the lower South Island of NZ. The two districts are rural^14^ and have a combined population of 61,000 dispersed over a wide geographical area (18,653 km^2^).^15^ The population is predominantly NZ European/Pākehā (86.5%), with the proportion who are Māori (6.4%) below the NZ average (16.5%).^15^ Gastroenterology services for Otago residents are provided by the Southern District Health Board (DHB)^16^ and delivered at Otago’s main hospital, Dunedin Hospital (a tertiary facility), by gastroenterologists and general surgeons, with an acute service available at all times. A rural endoscopy service is provided in the Central Otago District (Dunstan Hospital) with regular clinics organised for planned patients. Rural specialist clinics (pre-scheduled specialist visits) are also provided in the Queenstown-Lakes District (Lakes District Hospital). Patients who require urgent gastroenterology interventions are referred to specialists based in Dunedin Hospital. Access to IBD specialist care for people residing in the Central Otago and Queenstown-Lakes districts generally involves traveling from rural towns to Dunedin Hospital for initial appointments, screening tests and procedures, as well as follow-up appointments, if unable to attend a pre-scheduled clinic in Dunstan or Queenstown, closer to where they live. Patients may also contact the gastroenterology clinical nurse specialist directly for telephone advice.

### Design and sampling

The study utilised a qualitative exploratory design.^17^ It was part of a broader qualitative process evaluation undertaken alongside a randomized controlled trial (RCT) of teleconsulting for people living with IBD in the Central Otago and Queenstown-Lakes Districts (ACTRN12617000389303).^18^ A purposive sampling approach was used to recruit people living with IBD who were enrolled in the trial. A maximum variation strategy was used with the aim of ensuring breadth in terms of demographic characteristics (e.g., type of IBD, age, sex, ethnicity, distance from urban centre).

### Data collection

Semi-structured interviews were conducted by GN between October 2018 and February 2019. The interviews used a topic guide (Supplementary file 1) designed to elicit participants’ accounts in their own words. The topic guide was refined throughout the interviewing process to further investigate emerging themes. The interviews lasted one hour on average and were conducted at the participant’s home. Interviews were digitally-recorded and transcribed verbatim.

### Data analysis

We used a multi-stage thematic approach for qualitative data analysis,^19^ assisted by Atlas.ti software. All transcripts were independently read and coded by two experienced researchers (GN, LR). Data immersion occurred through repeated readings of the transcripts. An initial list of codes was generated iteratively through a first round of coding and discussed (GN, LR, TSt), with new codes being created as necessary. Codes were assigned to key sections of data to reflect the content. A subset of transcripts were also reviewed by TSt. This led to the development of a preliminary coding framework generated by regrouping codes with common features into emergent themes. The coding framework was discussed with the wider research team and further refined and validated through a second round of coding (GN, LR). Emerging themes were finally assigned to five overarching analytical constructs. The consolidated criteria for reporting qualitative research (COREQ)^20^ were used to inform reporting of the study findings (Supplementary File 2).

### Patient and public involvement

Patients or members of the general public were not involved in the design or conduct of this study.

## RESULTS

We interviewed 18 people living with IBD. The demographic characteristics of participants are shown in Table 1. Ten participants were in the RCT control group and eight were in the intervention group. The thematic findings describing peoples’ accounts of living with IBD are reflected in the following five analytical constructs: 1. Journey to confirming and accepting diagnosis; 2. Importance of the relationship with the health care team; 3. Support from others; 4. Learning how to manage IBD; and 5. Care at a distance - experiences of rurality. Illustrative quotes from participants are provided (Con# – control participant; Int# – intervention participant).

**Table 1.** Demographic characteristics of participants (n = 18)

|  |  |
| --- | --- |
| ***Type of IBD***<br>Ulcerative Colitis<br>Crohn's | 12<br>6 |
| ***Age***<br>30-39<br>40-49<br>50-59<br>60-69<br>70-79 | 4<br>5<br>5<br>1<br>3 |
| ***Sex***<br>Female<br>Male | 12<br>6 |
| ***Ethnicity***<br>NZ European / Pākehā | 18 |
| ***Travel distance between participants' rural residence and Dunedin Hospital***<br>Less than 150 km<br>150-200km<br>Greater than 200km | 0<br>7<br>11 |

### 1. Journey to confirming and accepting diagnosis

#### Pathways to confirming diagnosis

Confirming diagnosis was considered an important first step towards accepting having IBD. Participants shared two contrasting journeys of obtaining diagnosis. The first path was characterised by a long process. This was marked by recurrent sickness episodes where participants had been feeling unwell and dealing with various symptoms and underwent multiple health screening tests and sometimes even hospitalisation. In this instance, participants reported the initial diagnosis remained unclear for a long time despite multiple visits to the doctor. In addition, they had to cope with the burden of symptoms for a prolonged period which was distressing at times – stomach ache, blood in stool, losing weight, nausea, vomiting, low energy, and considerable impact on diet and daily routine. For these participants, diagnosis came as a relief and confirmation of a condition that they had been living with for a while.

> *“[It took] a long time, I was just in agony. I went to the [city] and was admitted to hospital with pain and they put me on IV antibiotics. They had done multiple tests, but no colonoscopy at that stage, and they sent me home, basically, just “put up with it”. [A while after that] I had to go back to my GP and then the [specialist] had requested a colonoscopy. [I had to travel to the city] for my colonoscopy and that’s when they found a lot of inflammation on my lower intestine.” (Int6)*

In contrast, the second path to diagnosis was portrayed as a more acute and short process in which participants suddenly became acutely unwell and required immediate medical attention, with some participants even requiring emergency surgery. In those circumstances, diagnosis was more of a shock.

> *“Diagnosis was a bit of a whirlwind. (…) I was shocked, it happened so fast. I didn’t really know what was happening. (…) I [remember going to the doctor] … within 40 minutes someone walked into the consultation room, lifted my top, drew a dot on my belly, threw me in an ambulance and performed emergency surgery at the local hospital. [The doctor] knew exactly what it was. ” (Int10)*

#### Developing acceptance of the condition (chronicity)

Travelling through life with IBD involved a long, and at times difficult, process of developing acceptance of the chronicity of the condition. Reflecting back on the initial stages following diagnosis, participants talked about the need for answers and seeking reassurance about the future. Acknowledging that the condition was something permanent that they would have to live with for the rest of their lives was central to the acceptance process.

> *“[A]s a patient, you’re looking for answers or for a cure. And I guess I’ve lived with this for a long time before I realised that potentially it’s not something that there’s a quick fix for. ” (Int3)*

The chronicity of the disease was particularly difficult to accept when going through more difficult periods involving unsettling flare-ups of symptoms.

> *“The biggest thing, of course, is that there’s no cure and it makes you wonder, ’Is it going to be like this [that uncomfortable], the way it is right now, forever?’” (Con6)*

Processing the news of diagnosis triggered a flood of emotions. Some participants reflected on feelings of grief, including denial, anger and sadness. Others talked about shame, the fear of judgement and stigma. For example, participants mentioned having to undergo repetitive invasive tests that can be shameful and dealing with symptoms that can be embarrassing.

> *“[If I] went to your house, I’d be like, I don’t really want to go the toilet because of loud intestinal gas (…). You’ve got to hold it in, just in case. (…) That can be embarrassing. Even a public toilet can be embarrassing” (Int10)*

Some participants had an awareness of the disease because of family history, and this was considered helpful in developing an understanding of the nature of the condition and the potential impact that it would have on daily life.

### 2. Importance of the relationship with the health care team

#### Relationship with the specialist team

The relationship with their specialist team (gastroenterologist and gastroenterology clinical nurse specialist) was considered a central component of participants’ journeys from receiving initial diagnosis to ongoing medical management and learning how to self-manage the condition. Building a strong relationship with the specialist in the initial stage following diagnosis was important to establish a good foundation for long-term management of the condition. Feeling listened to at specialist consultations was a key premise to building trustful connections.

> *“I don’t just feel like another IBD patient with them (specialist health care team).” (Con9)*

Participants’ main expectations of consultations with the specialist included obtaining clear information about the state of their condition and receiving guidance on medical management. Difficulty with understanding the specialist’s explanations or questions left unanswered had participants feeling “kept in the dark” at times. This was further nuanced by this participant explaining that information about the condition needs to be repeated multiple times as it can be a lot to take in at first, and understanding of it grows as you live with the condition over in time.

> *“I guess, just reiterating some of the information and telling people on more than one occasion, because you might hear something but you only pick up so much. And if that can be reiterated other times, then you (progressively develop) an understanding. (…) And the [Specialist] did explain this to me, you know, so he is giving me information, but I guess part of it is my understanding at the time that information was given to me and now that I’ve grown and learned more about it, some of that information would probably make more sense now than it probably did when he first told me.” (Int4)*

The specialist team’s openness to alternative ways of managing IBD was also considered important to many participants to foster open discussions about non-medical approaches that they wished to explore. Overall, relational continuity with the specialist team was viewed positively and allowed for a more in-depth and shared understanding of participants’ journeys to develop over time.

#### Relationship with the primary health care team

Participants emphasised the importance of local management of their condition by their general practitioner (GP) with whom most of them had long-established relationships.

> *“It’s been pretty well managed (locally). I have a good relationship with my GP (…). My GP was really involved when I was initially sick. Since then I am quite heavily involved with the specialist for my hepatitis (…) but as I stabilize it will be more GP management.” (Con1)*

Local management of IBD also prevented unnecessary travels to the city for health concerns that could be dealt with locally. For this to work best however, participants highlighted the need for good communication between their local GP and the specialist team in the city to ensure that decisions made on both ends are shared.

> *“(Communication between my specialist and my GP) is actually pretty good. My GP’s got the notes (from the specialist) and he reads them. (…) When I go and see him he’s read them. It’s like, (direct to the point).” (Con7)*

### 3. Support from others

Support from others was instrumental in enabling participants to progress through accepting the disease and developing confidence in everyday management of the condition.

#### Support from family and friends

Support from family and friends was considered critical in helping participants get through tough times at different stages of their illness. Participants shared reflections on how IBD impacts the whole family and not just the person with the diagnosis.

> *“It’s really daunting (…) and this is because it’s not just about me, it’s about the whole family.” (Con6)*

Support from significant others was also important to facilitate the implementation of various changes in daily routine, including diet.

> *“My husband had taken on board the gluten free diet and he does a lot of the cooking which is good. He was happy to incorporate that.” (Con15)*

Close friends were also part of the support network that helped with navigation through more difficult periods. Participants discussed how friends would bring fun and a little bit of banter at times where things were difficult, and this contributed to a positive outlook on life.

> *“My friends all know that I have this [IBD], so we treat it (with a bit of fun) now. (When we do activities, they would say): “Who’s going to flat with her, who’s going to be in the room with her, you know?” Because you have to, or else it just chews you up inside, and I’m not going to spend my life not going to concerts or going out with my girlfriends or going camping, because then, who’s the winner there? It’s not me, and not my family, and not my friends.” (Int2)*

#### Support from someone who also lives with the condition

Participants reflected on the support that they had received from people who live with the same condition. Many reported that it helped them with “normalising” the situation and allowed them to share common experiences and tips and tricks as they went through similar challenges. Having someone you could identify with provided reassurance.

> *“It wasn’t until I got home, and my wife started mentioning some names, like the man from the (local) shop, he had irritable bowel and he had a colostomy bag on. (…) (Next time I went to the shop) I start talking to him. He had the (colostomy bag) for a while and I was new to this, so he gave me a rundown of how things work. (…) He was a mentor, if that makes any sense.” (Int10)*

Support groups were not discussed at length but some participants were open to the idea of exchanging information with wider groups of people affected by the same condition as them. However, such groups were not always readily available in more isolated rural places.

### 4. Learning how to manage the condition

Another important aspect discussed by participants was learning how to progressively manage their condition and finding strategies that worked best for them in different circumstances. The following quote illustrates this well:

> *“[I]nitially I had that sort of naïve sense: “oh something’s wrong, the (specialist) is going to be able to fix it”. And then I learned that actually, it’s not so much about fixing it, it’s about managing it. (…) And finding what works for you (…).” (Int4)*

#### Developing an understanding of “what’s right for you”

Learning to manage the condition involved developing knowledge of treatments and familiarising yourself with the terminology surrounding them. This was considered particularly important to enable discussions about treatments with the specialist health care team. Making changes in your daily routine to help with managing symptoms, including identifying what you can and cannot eat, or arrangements around work and travel including locating public toilets when planning transportation, were seen as critical for finding a way of coping with the disease in the longer term. The learning process was often described as “trial and error” with participants testing out different strategies at different times to avoid deterioration of their condition.

> “*[T]here’s been a learning curve of what my triggers are, so I’ve sort of cut out coffee, I definitely don’t drink much wine these days, I cut out beer, I cut out gluten…” (Int4)*

This learning process also involved getting to know their limits and developing experiential knowledge that would allow them to build confidence in self-managing their condition.

> *“[Y]ou can go ahead and buy a steak … (and then realise) “I’m too bloated”. Your body can’t process it (…). Then you come home and you’re really tired, you just want to lie on the couch, and you feel anxious. You feel really uncomfortable (…) Now I can pick it up before it happens.” (Int10)*

#### Medical management and alternative approaches

Medication was presented as a central component of IBD management. Learning about medication was an important step towards controlling symptoms, but it had been challenging for many, particularly in the earlier stages following diagnosis.

> *“Medication. That’s the main thing. I take Colistin in the morning and PentasaI (…) I know if I wasn’t taken those I would have to go (to the toilet) more frequently. (The main thing) is definitely medication. I can’t get by without the medication.” (Con7)*

The use of technology was seen as an enabler to learning about medication and side effects, and it also provided easily accessible information about alternative strategies to cope with symptoms and IBD-related changes in their lives. As a complement to medication, many participants reported using different approaches to help with managing symptoms, including diet changes, acupuncture, and stress management programmes. Many participants considered those alternative measures as having a positive impact on the overall management of their condition.

> *“I have a good understanding (of my condition) and a good holistic view of what works for me and I have looked at alternatives so it certainty works for me having some diet changes and some other supplementary natural kind of medicines, managing stress, doing exercise and other things like that.” (Con1)*

#### Stress management in everyday life

Stress was identified as one of the main contributors to worsening of symptoms.

> *“[S]tress is the big thing, … I try not to be stressed out by things. (…) [I]t has a big part to play.” (Con3)*

Participants reported having to make significant adjustments in the way that they organise their lives, with one of the most prominent examples relating to how they manage work.

> *“[O]ver the years my workload has definitely picked up and my work is quite a “fight flight” kind of job, where you have constant interruptions, a lot of pressure, and I definitely felt over the years that (my work) contributed quite a lot to my symptoms, … and having flare ups depending on how stressful and stressed I was at work, so I think stress management for me is a real key.” (Int 4)*

Participants explained how long travel from rural towns to attend specialist appointments in the city, and transport more generally, could be stressful at times and require careful pre-planning. This included for example, identifying public toilets on route (e.g. via prior Google map searches or the use of mobile apps) and preparing snacks for the road to avoid having to buy food on the way.

Overall, a positive attitude to life transcended the discourse of participants with multiple examples of strength and resilience in the face of challenges. Participants shared reflections on how having a positive outlook on life influenced how they coped with the initial news of the diagnosis and adapted to the condition in time.

> *“That’s just something you live with, you battle yourself and you manage yourself in your own sort of way. (…) I’m a great believer in not putting the handles on the bath until you need them.” (Int3)*

### 5. Care at a distance - experiences of rurality

When describing their experiences of care, participants framed rurality as the inability to easily access specialist care. Living rurally had particular implications for ongoing management of IBD. One of the main challenges reported by participants was cost – cost of time and cost of mobilising resources due to long travel distances to get to Dunedin which sometimes required overnight accommodation, along with significant schedule disturbances.

> *“Living rurally means that I often have to go to the [urban centre] for treatment. (…) It costs me time to go back and forth. It costs me a hotel stay.” (Con5)*

Participants also shared reflections on privacy issues due to living in a small place where “everybody knows everybody“. Concerns were raised about having to undergo invasive tests at the local rural hospital where carers were sometimes neighbours and/or people the participants had known their entire life. The fear of access to personal information by people they knew also motivated some participants to seek care out of town.

> *“I travel to Dunedin for colonoscopies rather than having them at the rural hospital, it gives just a bit more privacy…It’s a pretty personal procedure. You really don’t want to see (people that you know) in the hallway.” (Con1)*

Managing IBD rurally involved challenges of access to specialist care, with perceptions of delayed referrals and concerns about disparities in specialist access compared to urban counterparts. Living far from the main health centre left some participants feeling like they were “falling through the cracks” and “forgotten about”.

> *“It has slowed things down, living in a small town (…). (…) being isolated in a small town has had implications like if I was in Dunedin (I would have seen) 50 doctors for different sort of things (…). Whereas here you don’t, you’ve only got your normal doctor in a small town (…).” (Int10)*

Feelings of isolation were apparent in the discourse of many participants when referring to living with IBD rurally.

> *“[I]t’s lonely living in the back country anyway. But living in the back country with a chronic illness…” (Con9)*

Receiving support locally from family and friends, as well as their local GP, were considered extremely important to get them through tough times.

## DISCUSSION

### Summary of findings

This study provides a rich understanding of the complex health journeys of people living with IBD and the challenges of managing the condition in rural areas of NZ. Five analytical constructs were identified: 1. Journey to confirming and accepting diagnosis; 2. Importance of the relationship with the health care team; 3. Support from others; 4. Learning how to manage IBD; and 5. Care at a distance - experiences of rurality. Pathways to confirming diagnosis involved two contrasting journeys: a long and slow process where diagnosis remained unclear for a prolonged period, and a more acute process where diagnosis typically came as a shock.

Central to the acceptance process was acknowledging the chronicity of the condition, which involved feelings of grief but also the fear of judgement and stigma. Building a strong relationship with the specialist was central to medical management, particularly in the initial stage following diagnosis. Good communication between the specialist and local GP was instrumental to local management of the condition. Support from significant others was critical to enable participants to progress through accepting the disease and developing confidence in everyday management of IBD. Sharing experiences with someone who also lives with IBD provided reassurance and helped with “normalising” the condition. Participants shared different strategies on how to manage IBD, describing a “trial and error” process of “finding what’s right” at different stages of the condition. Managing IBD rurally involved challenges of access to specialist care, with perceptions of delayed referrals and concerns about disparities in specialist access compared to urban counterparts. Rural living also had implications of costs - cost of time and cost of mobilising resources for long travels to the urban centre for treatments. Concerns about privacy issues were raised due to living in a small place where “everybody knows everybody“.

### Strengths and limitations

This qualitative interview study utilised purposive sampling to recruit a sample that was diverse in terms of the characteristics (type of IBD, age and sex) of people living with IBD rurally. Through the choice of our qualitative exploratory design we were able to obtain rich descriptions of people’s journeys at different stages of their disease, from confirming diagnosis through to managing IBD in their daily lives, including accounts of the importance of the relationship with the specialist and primary care teams, support from loved ones, helpful strategies to manage the condition, and aspects of rurality restricting service access to people living with IBD. This study was based in a single NZ health region, which provided rich context-specific findings that will benefit the local and regional health system. The findings may also be generalisable to other national health systems with large geographic areas and low population density.

We are cognisant of the timing of our research in the context of an ongoing and related trial and qualitative process evaluation subjecting participants to multiple contacts from different projects during the course of the present study. We also acknowledge that our participants comprised people who may have been more willing to share their journeys about IBD as they were already taking part in other studies and therefore their views may not fully reflect the wider range of experiences of people living with IBD rurally. The findings also only reflect the views of people living with IBD. To help inform service improvement, in future work it would be useful to seek the perspectives of the wider family and health care professionals. We were not able to recruit any ethnic group other than NZ European/Pākehā into this study. This may reflect both the population characteristics of the two Otago district catchment areas (predominantly NZ European)^15^ and the fact that IBD is less prevalent in Māori; future research focused on experiences for Māori is warranted.^3^

### Comparison with existing literature

There is a very limited NZ literature on living with IBD. The one previous NZ qualitative study focussed on four adolescents’ experiences of living with IBD.^21^ Our finding that the journey to confirming a diagnosis of IBD involved two contrasting paths: one a long process marked by recurrent symptoms and multiple health professional encounters, the other by becoming acutely unwell and requiring immediate medical assessment adds to the limited international literature in this area.^4 13^ Further research in this area could benefit from using the approach taken in early colorectal cancer diagnosis research in NZ^22^ and internationally^23^ which is to systematically explore the events and processes that occur from patients’ initial awareness of a bodily change to the start of treatment using the Models of Pathways to Treatment framework.^24^ The finding of the need to develop acceptance of the condition is also found in the international literature.^4 13 25 26^ Similarly, our findings of the importance of support from others (both from family and others who live with IBD) and the need to learn how to manage IBD have also been reported.^4 13 25^ Previous UK research has highlighted the need to adapt to living with IBD and that the concept of a “new normal” is core to adaptation – patients recover a sense of normality by achieving an equilibrium between their lives pre and post diagnosis.^25 26^ Our study also highlights the complex interplay of IBD symptoms, support from others and adaptation strategies which determine whether or not patients are able to achieve a “new normal.”

There is a limited international literature on how health care professionals help patients manage their IBD.^4 13 27 28^ One previous Swedish study has highlighted the importance of the need for members of specialist gastroenterology teams to treat patients with respect and mutual trust and to provide timely and clear information about management.^28^ The value of having a health care system which allows relational continuity of care^29^ with the specialist team has also been highlighted.^27 28^ Our study adds to this literature through identifying the need for the specialist team to be open to alternative ways of managing IBD so as to allow open discussions regarding non-medical approaches to management. Further, our study also highlights the importance of both relational and informational continuity of care^29^ with the primary health care team in terms of sharing care between a specialist team based in Dunedin and general practitioners based locally.

While there is Canadian evidence for significant rural/urban disparities in the quality of IBD care – rural IBD patients have lower rates of gastroenterologist physician visits, more hospitalizations, and greater rates of emergency department visits^9^ – no studies exploring rural patients’ perspectives on living with and managing IBD were identified. Our study identified two important barriers to accessing^30^ specialist IBD care rurally: ability to reach the specialist clinic in the urban centre and ability to cover the attendant costs (both direct and indirect) of travel. These barriers to access were also identified in a recent study of a different chronic disease (Chronic Obstructive Pulmonary Disease) set in the same region of NZ.^31^

### Implications for clinical practice and health policy

Patients living rurally with IBD value relational continuity of care with the gastroenterology specialist team which allows trust to develop and the provision of information and support at all stages of the patient’s journey: from receiving initial diagnosis through ongoing medical management during both remission and flare ups. For rural patients, however, this is often “care at a distance”, as attendance at the urban centre for treatment in person necessitates additional costs in terms of accommodation, fuel and the time taken for a lengthy commute.^32^ In order to address these access barriers there is a need to deliver and evaluate alternative methods of delivering gastroenterology care such as telehealth, online support, and remote outreach clinics.^10 11^ Telehealth, in particular, is increasingly used by NZ’s DHBs as a way of delivering virtual outpatient clinics across a range of clinical specialities.^33^ Another NZ development specific to IBD is the use of smartphone-based health applications as an alternative to outpatient visits for those with mild to moderate IBD.^34^ Further research is needed to determine if such virtual technologies lead to improvements in health outcomes as well as processes of care in patients living rurally.^12^

## CONCLUSIONS

This study provides a rich understanding of the complex health journeys of people living with IBD and the challenges of managing the condition in rural areas of NZ.

## Data Availability

Full de-identified interview transcripts will not be shared. Informed consent, in line with the approving ethics committee, only allows for the use of de-identified extracts within research reporting and writing, in order to maintain the privacy of participants based in a defined regional area and population, thus making their identification with full transcripts more likely.

## SUPPLEMENTARY FILES

Supplementary File 1: Interview Topic Guide. The semi-structured interview topic guide questions and probes. (.pdf)

Supplementary File 2: COREQ-32 reporting checklist. An assessment of the study reporting against the domains of the COREQ-32 reporting checklist for interviews and focus groups. (.pdf)

## ACKNOWLEDGEMENTS

We would like to acknowledge the study participants for their rich contributions to our understanding of the journeys of people living with IBD and the specific implications of rurality in terms of access to health care.

## CONTRIBUTORSHIP STATEMENT

TSt conceived and designed the study with input from LR, SD, TSu, AM, CH, FDN and MS. GN conducted the interviews. Data analysis was undertaken by LR and GN, with input from TSt and the wider research team. LR and TSt led the writing of the manuscript with input from GN. All authors reviewed and critiqued the manuscript and approved the final manuscript.

## ETHICS APPROVAL

Ethical approval was obtained from the University of Otago Human Ethics Committee (D18/145). Written informed consent was obtained from all participants.

